# Perceived disrespect and abuse among women delivering at a tertiary care center in Nepal

**DOI:** 10.1101/2021.01.23.21250363

**Authors:** Sabika Munikar, Mala Chalise, Ranjan Dhungana, Durga Laxmi Shrestha, Naresh Pratap KC, Animesh Dhungana, Robert B. Clark, Michael K. Visick, Kanchan Thapa

**Affiliations:** Post Basic Bachelor of Nursing Science Faculty, Om Health Campus, Kathmandu, Nepal; Independent Researcher from Kathmandu, Nepal; Helping Babies Breathe (HBB) Program, Safa Sunaulo Nepal, Kathmandu, Nepal; Ministry of Health and Population, Nepal; Department of Public Health, Brigham Young University, Provo, Utah, USA; Department of Pediatrics, University of Utah School of Medicine, Utah, USA; Central Department of Population Studies, Tribhuvan University, Kathmandu, Nepal

**Keywords:** Delivery, Disrespect and abuse, Labor, Maternal health services, Respectful maternity care, Midwives

## Abstract

**Background:** Of the children born every year in Nepal, 57.4% are delivered in health facilities. Disrespect and abuse of women during maternity care are problems that can significantly impact women’s willingness to seek out life-saving maternity care. However, evidence suggests ongoing disrespectful maternity care worldwide. This study aims to identify perceived disrespect and abuse during labor and delivery among postnatal women delivering at Bheri Hospital, Nepal.

**Methods:** A cross sectional study was conducted among 445 purposively selected women admitted in postnatal ward of Bheri Hospital, Nepal from February to March 2020. Ethical approval was obtained from Nepal Health Research Council. Informed written consent was obtained from each participant and a face-to-face interview was conducted for data collection. A semi-structured questionnaire consisting of demographic information and a pre-validated Respectful Maternity Care (RMC) tool was used. The information was then checked, coded, and entered in SPSS for descriptive and inferential analysis.

**Results:** In this study, the participants perceived very high friendly care, abuse-free care and discrimination-free care but moderate timely care only. Timely care was found to be significantly associated with age, ethnicity, occupation, monthly income, gravida, type of delivery, and complications. On multinomial regression, monthly income and type of delivery were the only factors found to be significant. Those mothers who had spontaneous vaginal delivery were 2.07 times more likely to have neutral RMC, and those who earn less than twenty thousand Nepalese rupees per month were likely to perceive high timely RMC.

**Conclusion:** This study concludes that disrespectful or abusive maternal care is not perceived among women delivering at Bheri Hospital in terms of friendly care, abuse-free care and non- discriminatory care. However, timely care is less reported. Appropriate interventions to provide timely care to delivering women must be instituted.

## Introduction

With Maternal Mortality Ratio (MMR) at 239 per 100,000 live births in 2016- higher than its South Asian neighbors- maternal mortality remains a formidable challenge in Nepal. Although the country has witnessed considerable decline in MMR by 55% from 1996 to 2016 [1], it still needs to go a long way to achieve the target of 70 per 100,000 live births as set out in the Sustainable Development Goals (SDGs) [2].

Ensuring access to quality skilled care before, during, and after childbirth is vital in reducing maternal mortality [3]. In low resource settings such as Nepal the lack of availability of skilled care services, mistreatment during childbirth, including abusive, neglectful, or disrespectful care may result in compromised quality [4]. Women have experienced disrespect and abuse (D &A) all over the world in various forms ranging from physical or verbal abuse, stigma or discrimination [4], detention of babies [4], being shouted at [5], threatening comments [5], withholding procedure related information and providing non-consented care [5]. For instance, a study in Ghana revealed that only a few clients were encouraged to ask questions and explained what to expect during labor [4]. Non-confidential care has also been reported [6], as identified in a study conducted in India. Similarly, evidences suggest that women have also experienced poor quality care in the form of restriction in their choice of birth position and movement, and restriction of liquid drinks during delivery [7].

Although a growing body of evidence paints a disturbing picture of women’s experience of care during pregnancy and child birth, health care providers justify such acts on the grounds of punishment for non-cooperation from women and good outcomes to babies[4]. Analyzed from the perspective of health service delivery system, difficult circumstances in health facilities under which maternity staffs work, system failures, and inadequate human resource management have been found as important reasons for D&A during delivery [8]. However, justifying disrespectful care and abuse based on these factors is a violation of women’s human rights.

RMC has been defined by World Health Organization (WHO) as “care organized for and provided to all women in a manner that maintains their dignity, privacy, and confidentiality, ensures freedom from harm and mistreatment, and enables informed choice, and continuous support during labor and childbirth” [9]. In this sense, RMC focuses on expanding safe motherhood beyond prevention of maternal mortality and morbidity to incorporate a human-rights based approach, including respect to women’s autonomy, dignity, choices, privacy and preferences [10]. RMC recognizes that all women need and deserve respectful care; and focuses on eliminating D&A during pregnancy and childbirth.

Despite the existing evidences that suggest D&A during childbirth presents considerable impediments to utilization of skilled birth care globally [9], only a few studies have been undertaken to understand the phenomena in Nepal. The majority of these studies have used a qualitative approach and only a limited number of studies have used a validated quantitative tool to measure the level of D&A at the point of service provision, out of which the greater number are based on health facilities in Kathmandu Valley.

This study aimed to identify perceived D&A during labor and deliveries among postnatal women admitted at a remote hospital and also determine the factors affecting RMC. Understanding women’s perspective of D&A during care is essential to identify factors that generate RMC in the health facility and subsequently in the provision of RMC as envisioned in The Right to Safe Motherhood and Reproductive Health Act of 2018.

## Methods

### Study Design, Study Setting and Sample Size

A cross-sectional study was done to identify the forms and associated risk factors of perceived disrespect and abuse among women delivering at Bheri Hospital, Nepal. With 5083 deliveries conducted in the year 2017-18 [11], Bheri Hospital in southwestern Nepal is a major referral center for emergency obstetric care services for three out of seven provinces (Lumbini Province, Karnali Province & Sudur Pachhim Province) of the country.

The sample size was calculated based on a study conducted in India, Ghana, and Kenya which depicted an overall prevalence of verbal abuse to be 16% across all countries [6]. Considering the prevalence of verbal abuse to be 16%, and level of significance to be 95%, the minimum sample size for the proposed study was calculated to be 237. However, we were able to collect the information from 445 women who delivered during the allocated period of data collection.

### Study Participants and Recruitment

Purposive sampling technique was used to interview postnatal women admitted at the postnatal ward of Bheri Hospital, within 24 hours of delivery. Those who were unwilling to participate in the study, couldn’t understand and/or speak the Nepali language or had a stillbirth or macerated birth during delivery were excluded from the study.

### Data Collection

Face-to-face interview technique was used to collect data. Each interview lasted for approximately 20 minutes and conducted in the Nepali language. The data were collected from February to March 2020. Informed written consent was obtained before data collection. Data were collected from women in postnatal ward within 24 hours of delivery to avoid recall bias.

A semi-structured questionnaire, divided into two parts, was used as the tool for data collection. The first part included questions relating to socio-demographic factors and obstetric history, details of which are presented in Table 1. The second part of the questionnaire was based on a validated RMC tool with 15 items used to measure women’s perception regarding RMC. The tool has four dimensions: friendly care, abuse-free care, timely care and non-discriminatory care consisting of 7, 3, 3 and 2 items, respectively [12]. The construct validity of the scale is confirmed by the high average factor loading of the four components ranging from 0.76 to 0.82 and a low correlation between the components. The scale has adequate reliability with α = 0.845[13]. The instrument was translated into the Nepali language and validated by a Nepali language expert. The content validity of the instrument was established by consultation with subject experts. Consistency of the tool was checked by pre-testing among 10% of women delivering at Bheri Hospital which was not included in the final study sample.

**Table 1.** Distribution of variables in the study

|  |  |
| --- | --- |
| **Part I-** Socio-demographic characteristics and obstetric history | **Demographic Information:** Age of Mother, Ethnicity, Religion, Type of Family, Educational Status, Educational Level, Occupation, Spouse’s Educational Status, and Spouse’s Occupation |
|  | **Obstetric History:** Gravida, Para, Week of Gestation, Type of Delivery, and Complication During Labor |
| **Part II-** Perceived Disrespect & Abuse <sup>12</sup> | **Friendly Care, Abuse-free Care, Timely Care, and Discrimination-free Care** |

### Data Analysis

The collected data was checked, organized and coded, and entered into Microsoft excel and then exported to SPSS (Statistical Package for the Social Sciences) 17.0 version for analysis. The data were analyzed by using descriptive statistics like frequency, percentage, mean, standard deviation, and inferential statistics: chi-square test, and multinomial logistic regression.

Mean score (M) of four broad components i.e. friendly care, timely care, abuse free care, and non- discriminatory care was used to describe the level of respectful maternity care experienced by the postpartum women during childbirth in each component separately. To determine the participant’s degree of respectful maternity care, the following Likert- range conversion and qualitative interpretation were used: 4.20- 5.00-Very High, 3.4- 4.19 -High, 2.60- 3.3 - Moderate, 1.80- 2.5 -Low and 1- 1.78 Very Low [12].

### Ethical Approval

Ethical approval was obtained from the Nepal Health Research Council (Ref #1953, 18 March 2020). Written permission from Bheri Hospital administration was also obtained. Informed written consent was obtained from the respondents. Participants were also assured that their participation/non-participation would have no bearing on their treatment. Confidentiality of the participants was maintained by assigning unique identification code to each participant.

## Results

Table 2 depicts the socio-demographic information of the participants. The majority of them (74.4%) were aged 20- 30 years, belonged to the Janjati ethnic group (46.1%), and most of them (91.2%) followed the Hindu religion. The majority (86.7%) were educated. However, almost half (50.8%) were unemployed. Approximately fifty-three percent of respondents were from joint or extended family. Regarding the spouse’s background, most of them (91.7%) were educated and were involved in a non-formal occupation (71.5%).

**Table 2.**
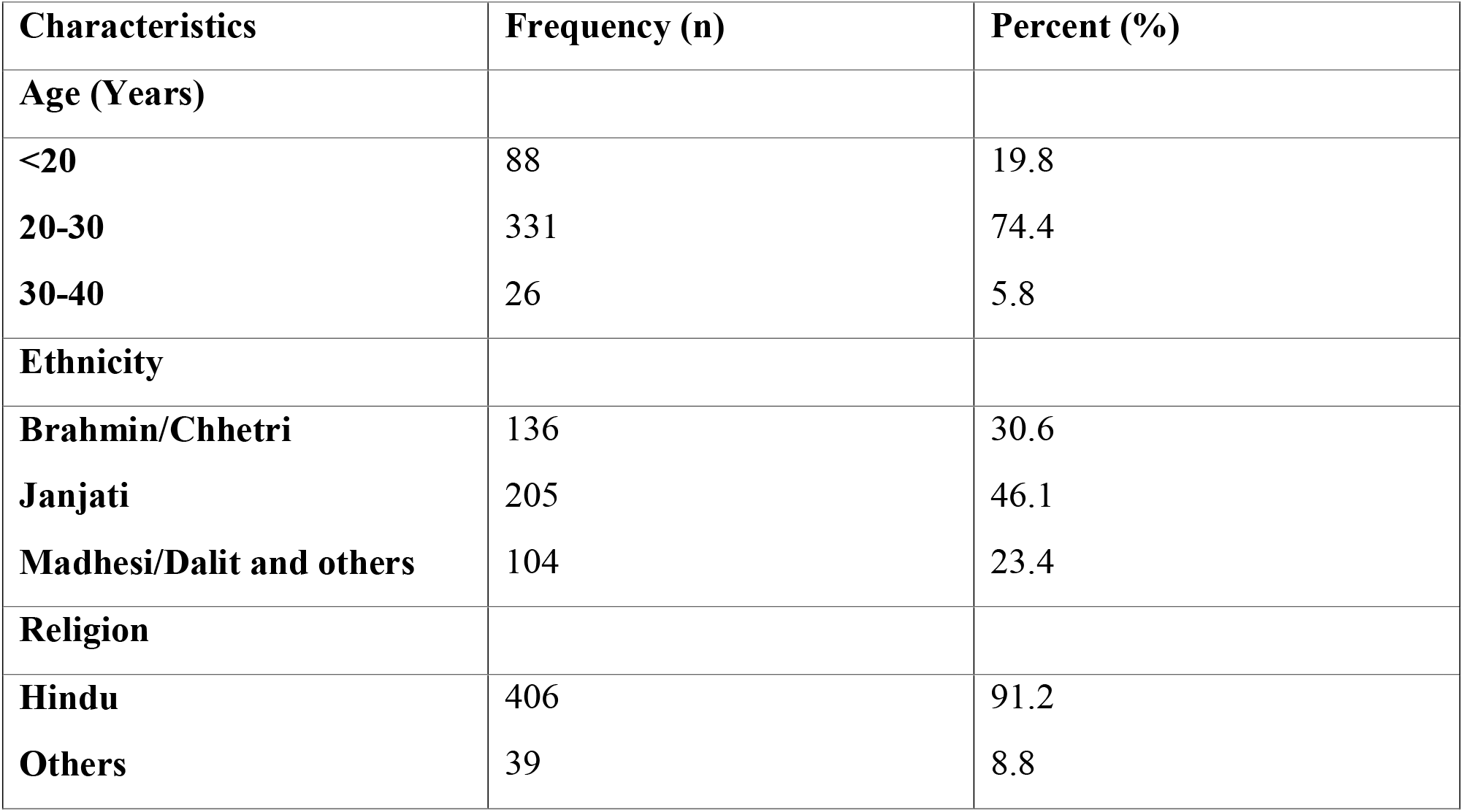
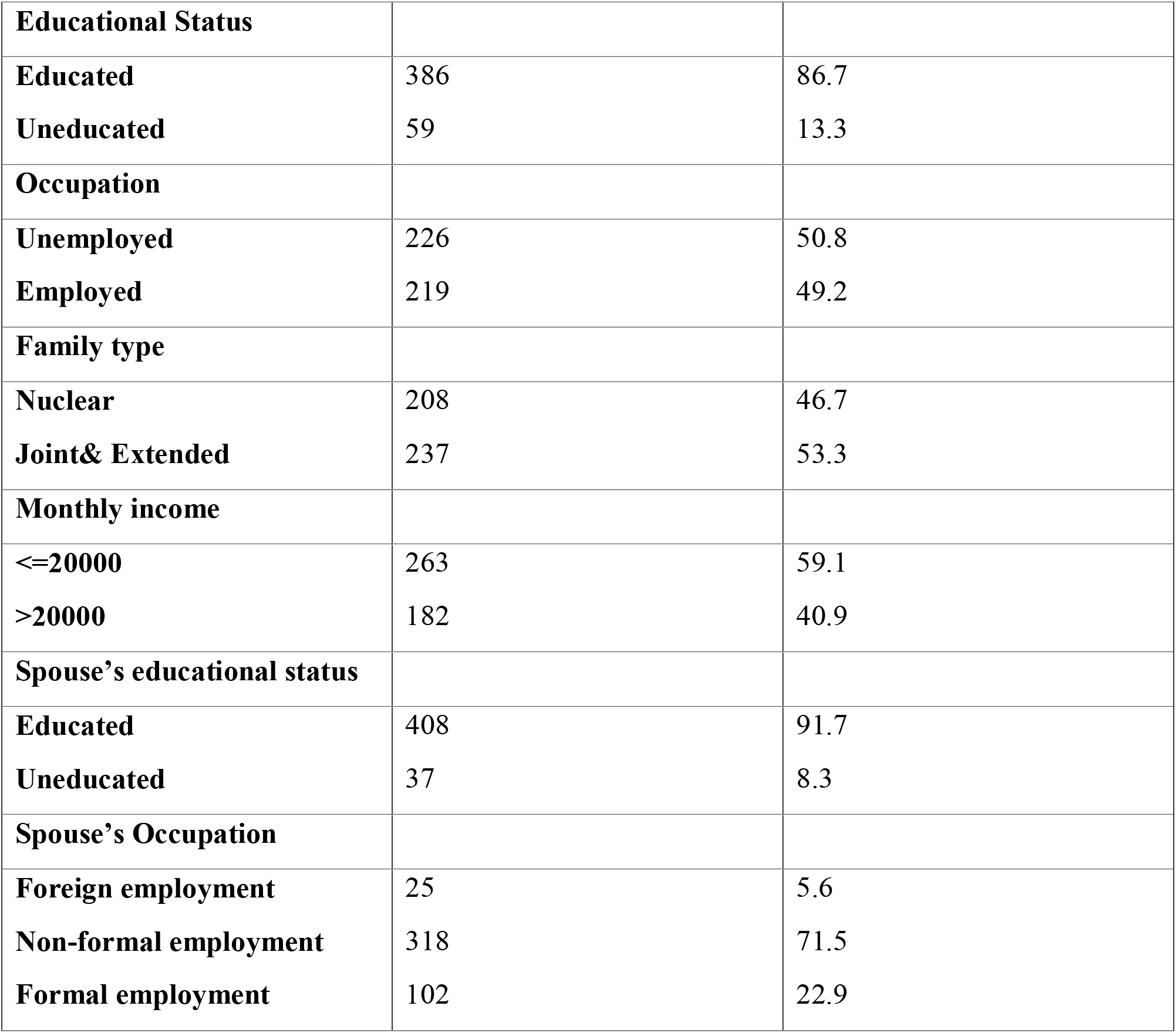
Distribution of participants according to socio-demographic information (n= 445)

| Characteristics | Frequency (n) | Percent (%) |
| --- | --- | --- |
| **Age (Years)** |  |  |
| <20 | 88 | 19.8 |
| 20-30 | 331 | 74.4 |
| 30-40 | 26 | 5.8 |
| **Ethnicity** |  |  |
| Brahmin/Chhetri | 136 | 30.6 |
| Janjati | 205 | 46.1 |
| Madhesi/Dalit and others | 104 | 23.4 |
| **Religion** |  |  |
| Hindu | 406 | 91.2 |
| Others | 39 | 8.8 |
| **Educational Status** |  |  |
| **Educated** | 386 | 86.7 |
| **Uneducated** | 59 | 13.3 |
| **Occupation** |  |  |
| **Unemployed** | 226 | 50.8 |
| **Employed** | 219 | 49.2 |
| **Family type** |  |  |
| **Nuclear** | 208 | 46.7 |
| **Joint& Extended** | 237 | 53.3 |
| **Monthly income** |  |  |
| **<=20000** | 263 | 59.1 |
| **>20000** | 182 | 40.9 |
| **Spouse's educational status** |  |  |
| **Educated** | 408 | 91.7 |
| **Uneducated** | 37 | 8.3 |
| **Spouse's Occupation** |  |  |
| **Foreign employment** | 25 | 5.6 |
| **Non-formal employment** | 318 | 71.5 |
| **Formal employment** | 102 | 22.9 |

Table 3 illustrates the obstetric history of participants. The majority of participants had less than two gravidae (75.7%), were multiparous (64.7%) and most of them (95.7%) had term pregnancy. Almost half of the participants (51.5%) delivered via spontaneous vaginal delivery (SVD) while remaining delivered via augmented labor and/or lower section cesarean section. One third of respondents (33.3%) had complications during labor.

**Table 3.** Distribution of participants according to obstetric history (n=445)

| **Characteristics** | **Frequency (n)** | **Percent (%)** |
| --- | --- | --- |
| **Gravida** |  |  |
| **<=2** | 337 | 75.7 |
| **>2** | 108 | 24.3 |
| **Para** |  |  |
| **Primi para** | 157 | 35.3 |
| **Multipara** | 288 | 64.7 |
| **Week of gestation** |  |  |
| **Preterm** | 19 | 4.3 |
| **Term** | 426 | 95.7 |
| **Type of delivery** |  |  |
| **Spontaneous vaginal delivery** | 229 | 51.5 |
| **Augmented** | 63 | 14.2 |
| **Lower section cesarean section** | 153 | 34.4 |
| **Complications during labor** |  |  |
| **Yes** | 148 | 33.3 |
| **No** | 148 | 66.7 |

Table 4 shows the perception of participants regarding RMC on a 5 points Likert scale. The components of RMC are presented in four broad categories of Friendly Care, Abuse-free Care, Timely Care, and Non-discriminatory Care.

**Table 4.** Participant reports of RMC (n=445)

| **Components of RMC** | **SD**<br>**f (%)** | **D**<br>**f (%)** | **N**<br>**f (%)** | **A**<br>**f (%)** | **SA**<br>**f (%)** | **Mean**<br>**(SE)** | **CM**<br>**(SE)** |
| --- | --- | --- | --- | --- | --- | --- | --- |
| **Friendly care: The health worker/s** |  |  |  |  |  |  |  |
| **Cared with a kind approach** | 3<br>(0.7) | 1<br>(0.2) | 1<br>(0.2) | 141<br>(31.7) | 299<br>(67.2) | 4.64<br>(.027) | 4.42<br>(.028) |
| **Treated in a friendly manner** | 10<br>(2.2) | 2<br>(0.4) | 27<br>(6.1) | 185<br>(41.6) | 221<br>(49.7) | 4.36<br>(.038) |  |
| **Talked positively about pain and relief** | 5<br>(1.1) | 4<br>(0.9) | 14<br>(3.1) | 200<br>(44.9) | 222<br>(49.9) | 4.42<br>(.033) |  |
| **Showed his/her concern and empathy** | - | 1<br>(0.2) | 38<br>(8.5) | 187<br>(42) | 219<br>(49.2) | 4.40<br>(.031) |  |
| **Treated with respect as an individual** | 1<br>(0.2) | 6<br>(1.3) | 66<br>(14.8) | 181<br>(40.7) | 191<br>(42.9) | 4.25<br>(.036) |  |
| **Spoke in a language that I could understand** | 18<br>(4) | 29<br>(6.5) | 24<br>(5.4) | 174<br>(39.1) | 200<br>(44.9) | 4.14<br>(.05) |  |
| **Called me by my name** | 113<br>(25.4) | 97<br>(21.8) | 46<br>(10.3) | 130<br>(29.2) | 59<br>(13.3) | 2.83<br>(.068) |  |
| **Abuse free care:** |  |  |  |  |  |  |  |
| **The health worker/s** |  |  |  |  |  |  |  |
| **Responded to my needs whether or not I asked** | 51<br>(11.5) | 94<br>(21.1) | 89<br>(20.0) | 90<br>(20.2) | 121<br>(27.2) | 3.31<br>(.065) | 4.29<br>(.044) |
| **Slapped me during delivery for different reasons (R)** | 267<br>(60.0) | 121<br>(27.2) | 32<br>(7.2) | 19<br>(4.3) | 6<br>(1.3) | 4.40<br>(0.43) |  |
| **Shouted at me because I hadn't done what I was told to do (R)** | 249<br>(56.0) | 125<br>(28.1) | 50<br>(11.2) | 17<br>(3.8) | 4<br>(0.9) | 4.34<br>(.042) |  |
| **Timely Care** |  |  |  |  |  |  |  |
| **Kept waiting for a long time before receiving service (r)** | 98<br>(22.0) | 117<br>(26.3) | 106<br>(23.8) | 78<br>(17.5) | 46<br>(10.3) | 3.32<br>(.061) | 3.10<br>(.057) |
| **Allowed to practice cultural rituals in the facility** | 169<br>(38.0) | 107<br>(24.0) | 87<br>(19.6) | 19<br>(4.3) | 63<br>(14.2) | 2.33<br>(.066) |  |
| **Service provision was delayed due to the health facility's internal problems (r)** | 172<br>(38.7) | 102<br>(22.9) | 88<br>(19.8) | 51<br>(11.5) | 32<br>(7.2) | 3.74<br>(.060) |  |
| **Discrimination- free care: Some of the health workers** |  |  |  |  |  |  |  |
| **Did not treat me well because of my personal attributes (R)** | 330<br>(74.2) | 89<br>(20.0) | 11<br>(2.5) | 9<br>(2.0) | 6<br>(1.3) | 4.64<br>(.036) | 4.67<br>(.032) |
| **Insulted me and my companions due to my personal attributes (R)** | 316<br>(71.0) | 112<br>(25.2) | 4<br>(0.9) | 5<br>(1.1) | 8<br>(1.8) | 4.62<br>(.035) |  |

SA- Strongly agree, A- Agree, N- Neutral/ Indifferent, D- Disagree, SD- Strongly disagree
SE- Standard error, CM- Cumulative mean

Regarding Friendly Care, very few respondents disagreed that the health workers cared for them with kind approach (0.9%), treated them in a friendly manner (2.6%), talked positively about the pain and relief measures (2.0%), showed concern and empathy (0.2%), treated them with respect as an individual (1.5%), and spoke in understandable language (10.5%). Of note, almost half of the participants (47.2%) disagreed on being called by their name.

The table also presents the perception of participants towards Abuse-free Care. Nearly 32.6% disagreed with the statement that health workers responded to their needs whether or not asked. Also, 5.6% reported being slapped during delivery for different reasons, and a similar number of participants (4.7%) reported being shouted at for not doing what they were told to do.

Regarding Timely Care, more than a quarter of participants (27.8%) agreed to being kept waiting for a long time before receiving care, but a higher number of participants were not allowed to practice cultural rituals (62%). Some agreed that service was delayed due to health facility’s internal problems (18.7%).

Perception towards Discrimination-free Care shows that few respondents (3.3%) agreed that the health workers did not treat them well because of personal attributes. Also, 2.9% of participants agreed that some health workers insulted them and their companions due to personal attributes.

The mean score shows that the participants’ perceptions of Non-discrimination Care (4.67), Friendly Care (4.42) and Abuse-free Care (4.29) were very high, whereas perception of Timely Care (3.10) was comparatively moderate.

Table 4 presents the findings of the association between selected demographic and obstetric characteristics and the Friendly Care component of RMC. This table shows that there is a significant association between timely care and monthly income (p<0.05), gravida (p<0.05), para (p=0.004), and week of gestation (p=0.026).

Table 6 reveals the association between Abuse-free Care and selected demographic and obstetric characteristics. This table shows that there is a significant association between Abuse-free Care and spouse’s occupation (p=0.002), para (p=0.010), type of delivery (p<0.05).

**Table 5.** Distribution of Friendly RMC by different socio-demographic and obstetric characteristics (n=445)

| Characteristics | Friendly care | | | $\chi^2$ | p-value |
| --- | --- | --- | --- | --- | --- |
|  | Moderate RMC | High RMC | Very high RMC |  |  |

|  | **f (%)** | **f (%)** | **f (%)** |  |  |
| --- | --- | --- | --- | --- | --- |
| **Age** |  |  |  |  |  |
| **<20** | 1 (1.1) | 41 (46.6) | 46 (52.3) | 8.958<sub>a</sub> | .062 |
| **20-30** | 24 (7.3) | 149(45.0) | 158 (47.7) |  |  |
| **30-40** | 1 (3.8) | 16 (61.5) | 9 (34.6) |  |  |
| **Ethnicity** |  |  |  |  |  |
| **Brahmin/Chhetri** | 10 (7.4) | 58 (42.6) | 68 (50.0) | 6.754 | .149 |
| **Janjati** | 8 (3.9) | 92 (44.9) | 105 (51.2) |  |  |
| **Madhesi/Dalit & others** | 8 (7.7) | 56 (53.8) | 40 (38.5) |  |  |
| **Educational Status** |  |  |  |  |  |
| **Educated** | 25 (6.5) | 177(45.9) | 184 (47.7) | 2.817<sub>a</sub> | .244 |
| **Uneducated** | 1 (1.7) | 29 (49.2) | 29 (49.2) |  |  |
| **Occupation** |  |  |  |  |  |
| **Unemployed** | 9 (4.0) | 115(50.9) | 102 (45.1) | 5.529 | .063 |
| **Employed** | 17 (7.8) | 91 (416) | 111 (50.7) |  |  |
| **Monthly income** |  |  |  |  |  |
| **<=20000** | 5 (1.9) | 126(47.9) | 132 (50.2) | 18.188 | **.000** |
| **>20000** | 21 (11.5) | 80 (44.0) | 81 (44.5) |  |  |
| **Spouse's Occupation** |  |  |  |  |  |
| **Foreign** | 0(.0) | 13(52.0) | 12(48.0) | 3.605<sub>a</sub> | .462 |
| **Non-formal** | 21(6.6) | 146(45.9) | 151(47.5) |  |  |
| **Formal employment** | 5(4.9) | 47(46.1) | 50(49.0) |  |  |
| **Gravida** |  |  |  |  |  |
| **<=2** | 7 (2.1) | 146(43.3) | 184 (54.6) | 49.498 | **.000** |
| **>2** | 19 (17.6) | 60 (55.6) | 29 (26.9) |  |  |
| **Para** |  |  |  |  |  |
| **Primi para** | 2 (1.3) | 83 (52.9) | 72 (45.9) | 11.135 | **.004** |
| **Multi para** | 24 (8.3) | 123(42.7) | 141 (49.0) |  |  |
| **Week of gestation** |  |  |  |  |  |
| **Preterm** | 0 | 14 (73.7) | 5 (26.3) | 7.302<sub>a</sub> | **.026** |
| **Term** | 26 (6.1) | 192(45.1) | 208 (48.8) |  |  |
| **Type of delivery** |  |  |  |  |  |
| **SVD** | 10 (4.4) | 100(43.7) | 119 (52.0) | 6.112<sub>a</sub> | .191 |
| **Augmented** | 6 (9.5) | 34 (54.0) | 23 (36.5) |  |  |
| **LSCS** | 10 (6.5) | 72 (47.1) | 71 (46.4) |  |  |
| **Complications** |  |  |  |  |  |
| **Yes** | 7 (4.7) | 69 (46.6) | 72 (48.6) | 0.504 | .777 |
| **No** | 19 (6.4) | 137(46.1) | 141 (47.5) |  |  |

<sub>a</sub> – likelihood ratio

**Table 6.**
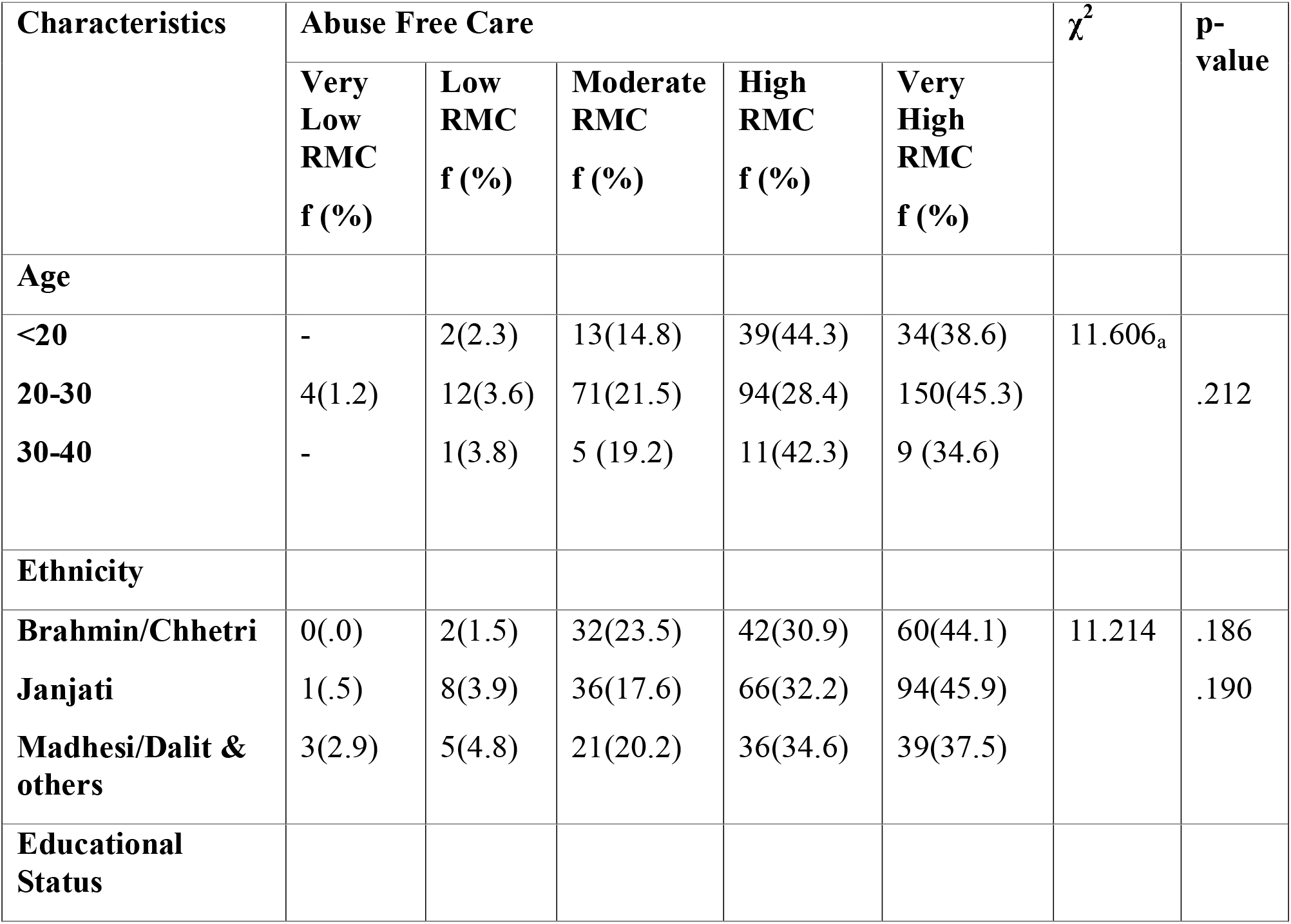
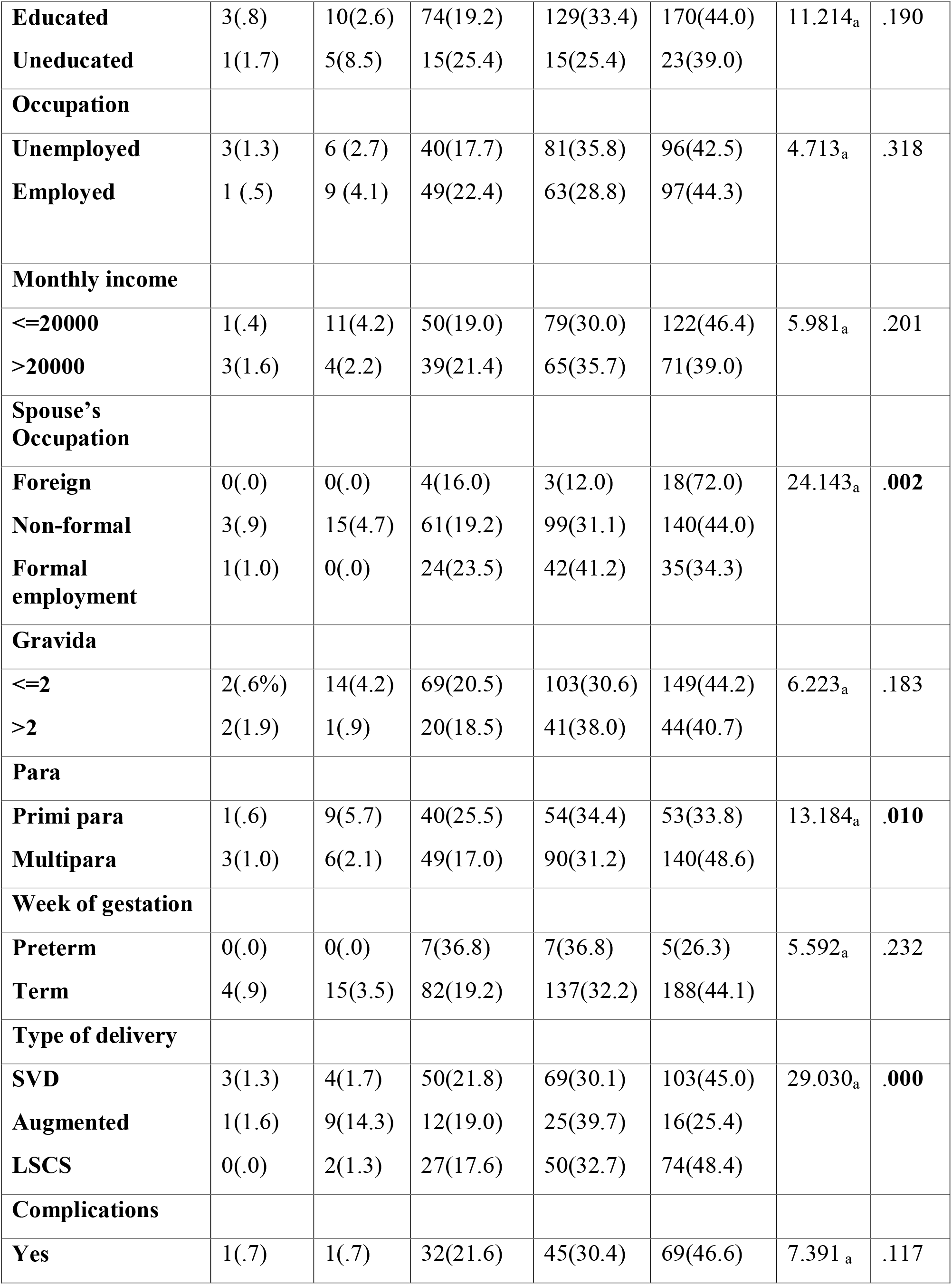
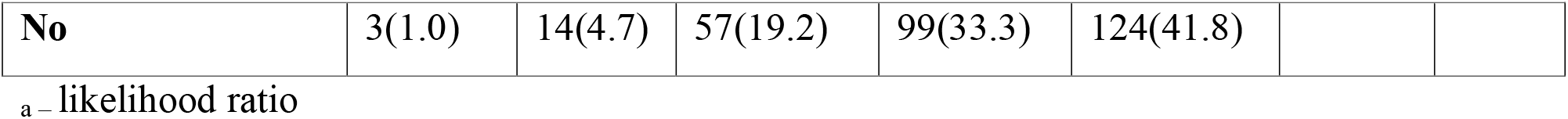
Distribution of Abuse-free RMC by different socio-demographic and obstetric characteristics (n=445)

Table 7 shows the association between Timely Care and selected characteristics which reveals that there is a significant association between Timely Care and age (P<0.05), ethnicity (p=0.002), occupation (p=0.001), monthly income (p<0.05), gravida (p<0.05), type of delivery (p=0.002), and complications (p=0.002). However, there is no significant association between Timely Care and educational status, spouse’s occupation, or week of gestation (P>0.05).

**Table 7.** Distribution of Timely RMC by different socio-demographic and obstetric characteristics (n=445)

| Characteristics | Timely Care | | | | | $\chi^2$ | p-value |
| --- | --- | --- | --- | --- | --- | --- | --- |
|  | Very Low RMC | Low RMC | Moderate RMC | High RMC | Very high RMC |  |  |
| **Age** |  |  |  |  |  |  |  |
| **<20** | 0(.0) | 8(9.1) | 37(42.0) | 25(28.4) | 18(20.5) | 55.822a | .000 |
| **20-30** | 38(11.5) | 33(10.0) | 179(54.1) | 73(22.1) | 8(2.4) | 54.618 | .000 |
| **30-40** | 3(11.5) | 1(3.8 0 | 13(50.0) | 9(34.6) | 0(.0) |  |  |
| **Ethnicity** |  |  |  |  |  |  |  |
| **Brahmin/Chhetri** | 19(14.0) | 13(9.6) | 75(55.1) | 29(21.3) | 0(.0) | 24.939a | .002 |
| **Janjati** | 12(5.9) | 17(8.3) | 101(49.3) | 53(25.9) | 22(10.7) |  |  |
| **Madhesi/Dalit & others** | 10(9.6) | 12(11.5) | 53(51.0) | 25(24.0) | 4(3.8) |  |  |
| **Educational Status** |  |  |  |  |  |  |  |
| **Educated** | 38(9.8) | 39(10.1) | 198(51.3) | 90(23.3) | 21(5.4) | 4.089a | .394 |
| **Uneducated** | 3(5.1) | 3(5.1) | 31(52.5) | 17(28.8) | 5(8.5) | 4.420 | .352 |
| **Occupation** |  |  |  |  |  |  |  |
| **Unemployed** | 10(4.4) | 29(12.8) | 113(50.0) | 61(27.0) | 13(5.8) | 18.888a | .001 |
| **Employed** | 31(14.2) | 13(5.9) | 116(53.0) | 46(21.0) | 13(5.9) |  |  |
| **Monthly income** |  |  |  |  |  |  |  |
| **≤20000** | 18(6.8) | 23(8.7) | 129(49.0) | 68(25.9) | 25(9.5) | 20.616a | .000 |
| **>20000** | 23(12.6) | 19910.4) | 100(54.9) | 39(21.4) | 1(.5) |  |  |
| **Spouse's Occupation** |  |  |  |  |  |  |  |
| **Foreign** | 6(24.0) | 2(8.0) | 13(2.0) | 4(16.0) | 0(.0) | 13.692a | .090 |
| **Nonformal** | 25(7.9) | 31(9.7) | 159(50.0) | 79(24.8) | 24(7.5) | 14.218 | .076 |
| **Formal employment** | 10(9.8) | 9(8.8) | 57(55.9) | 24(23.5) | 2(2.0) |  |  |
| **Gravida** |  |  |  |  |  |  |  |
| **<=2** | 17(5.0) | 32(9.5) | 185(54.9) | 85(25.2) | 18(5.3) | 30.782a | **.000** |
| **>2** | 24(22.2) | 10(9.3) | 44(40.7) | 22(20.4) | 8(7.4) |  |  |
| **Para** |  |  |  |  |  |  |  |
| **Primi para** | 8(5.1) | 18(11.5) | 87(55.4) | 39(24.8) | 5(3.2) | 9.255a | .055 |
| **Multi para** | 33(11.5) | 24(8.3) | 142(49.3) | 68(23.6) | 21(7.3) |  |  |
| **Week of gestation** |  |  |  |  |  |  |  |
| **Preterm** | 2(10.5) | 0(.0) | 10(52.6) | 5(26.3) | 2(10.5) | 2.703a | .609 |
| **Term** | 39(9.2) | 42(9.9) | 219(51.4) | 102(23.9) | 24(5.6) | 4.355 | .360 |
| **Type of delivery** |  |  |  |  |  |  |  |
| **SVD** | 20(8.7) | 14(6.1) | 129(56.3) | 56(24.5) | 10(4.4) | 22.359a | .004 |
| **Augmented** | 10(15.9) | 7(11.1) | 32(50.8) | 14(22.2) | 0(.0) | 24.612 | **.002** |
| **LSCS** | 11(7.2) | 21(13.7) | 68(44.4) | 37(24.2) | 16(10.5) |  |  |
| **Complication** |  |  |  |  |  |  |  |
| **Yes** | 10(6.8) | 22(14.9) | 68(45.9) | 33(22.3) | 15(10.1) | 16.957a | **.002** |
| **No** | 31(10.4) | 20(6.7) | 161(54.2) | 74(24.9) | 11(3.7) |  |  |
| **No. of living child** |  |  |  |  |  |  |  |
| **≤2** | 22(5.8) | 37(9.8) | 204(54.0) | 92(24.3) | 23(6.1) | 35.113a | .000 |
| **>2** | 19(28.4) | 5(7.5) | 25(37.3) | 15(22.4) | 3(4.5) | 26.558 | .000 |

a\_ likelihood ratio

As mentioned in Table 4, among the four components, women’s perception of Timely Care was found to be moderate whereas other dimensions of RMC were perceived very high. To determine the factors resulting in moderate perception of Timely Care, multinomial logistic regression was done. Table 8 shows that those who had SVD were 2.07 times as likely to have neutral RMC for Timely Care. Similarly, those who earn less than twenty thousand Nepalese Rupees per month were 2.36 (1.30-4.23) times as likely to have high Timely RMC in Nepal. We did not observe any significant effects between gravida, complication during the delivery, and the number of living children (P>0.05).

**Table 8.** Associated factors for Timely RMC during delivery (n=445)

| Predictors | Neutral RMC |  | High RMC |  |
| --- | --- | --- | --- | --- |
|  | OR( CI) | P-Value | OR(CI) | P-Value |
| Type of Delivery |  |  |  |  |
| SVD | 2.07(1.15-3.72) | 0.01 | 1.38(0.74-2.59) | 0.30 |
| Augmented | 0.99(0.47-2.10) | 0.99 | 0.56(0.23-1.36) | 0.18 |
| LSCS | 0<sup>b</sup> | - | 0<sup>b</sup> | - |
| Gravida |  |  |  |  |
| Primi | 2.28(0.92-6.29) | 0.07 | 2.20(0.82-5.90) | 0.12 |
| Multi | 0<sup>b</sup> |  | 0<sup>b</sup> | - |
| Monthly income (NPR) |  |  |  |  |
| ≤20000 | 1.33(0.78-2.24) | 0.289 | 2.36(1.3-4.23) | 0.004 |
| >20000 | 0<sup>b</sup> | - | 0<sup>b</sup> |  |
| Complication during delivery |  |  |  |  |
| Yes | 0.81(0.43-1.53) | 0.52 | 0.63(0.31-1.27) | 0.63 |
| No | 0<sup>b</sup> | - | 0<sup>b</sup> |  |
| Living Children |  |  |  |  |
| ≤2 | 1.86(0.74-2.59) | 0.048 | 1.52(0.52-4.39) | 0.43 |
| ≥2 | 0<sup>b</sup> | - | 0<sup>b</sup> | - |

Model fitting information= Chi-square= 3.68,df=12 p-value=0.000, Pseudo R<sup>2</sup> Nagelkerke=0.096
a. The reference category is 1.00 Low RMC, b This parameter set zero because it is redundant,

## Discussion

This study aimed to identify perceived Disrespect & Abuse and its associated factors during labor and delivery among postnatal women at a busy referral hospital in western Nepal. D&A are evaluated based on four different dimensions of RMC i.e. Friendly Care, Abuse-free Care, Timely Care and Discrimination-free Care. Very high degree of Friendly Care, Abuse-free Care and Discrimination-free Care was identified, however, only moderate Timely Care was perceived by the participants which is in contrast to the study in Egypt where only Discrimination-free Care was perceived to be high and other dimensions to be moderate [12].

The reason for high rating of Abuse-free Care in this study could be normalization of the abuse in the health care setting [14], where delivering women think that it is normal to be abused physically and/or verbally for better labor outcomes. Also, despite the knowledge of principles of RMC among health care providers, this knowledge may not translate to an improvement in actual respectful care at the bedside [14].

Most of the women (91.3%) perceived that they were treated in a friendly manner which is consistent with a direct observation of RMC in health facilities of five countries in East and Southern Africa (86%) [7]. Talking positively about the pain and relief measures was one of the components of Friendly Care where very few respondents (2.0%) disagreed with the statement. The reasons for not addressing pain may be due to the lack of availability of a doctor [14] and/or the shortage of health workers persistent in the country [15]. The shortage, however, might have been more pronounced at the time of data collection due to the ongoing staff adjustment process undertaken as part of implementing Federalism in the country [16].

WHO recommends communication between maternity care providers and women in labor, using simple and culturally acceptable methods [17]. Evidence suggests that language barrier is a critical factor that hinders effective communication and can also pose considerable risk to patient safety and quality of care [18]. More than two third of the participants (84%) in this study confirmed that the health worker spoke in a language understandable to them. A study conducted in Egypt found that nearly 61% of the health workers did not communicate in an understandable language. This suggests that language barrier was less common in our context. To ensure respectful attitude and supportive environment during delivery, it is required to continue emphasizing the importance of health care provider-client communication and client-centered care [19].

One of the critical elements affecting patients’ perception of RMC is the way in which a patient is addressed by a name of her/his preference. Patients’ preferred mode of address by healthcare workers, to large extent, is influenced by ethnic and cultural factors. For instance, a study on non-English speaking Australians shows that patients preferred to be called by their informal name [20]. On the other hand, patients in countries like Iran [21] and Israel [22] preferred formal address by title and surname. In this study, 47.2% respondents agreed that they were called by their preferred name, which is similar to the study from Egypt [12]. However, with limited evidence on Nepalese patients’ preference of address by healthcare workers, the present study is unable to provide contextual interpretation of the figure. Therefore, we recommend further study on Nepalese patients’ preferred mode of address by healthcare workers.

Neglect or abandonment during labor and delivery has been reported in varying degrees in countries like Kenya (14.3%) [23] and Tanzania (3.45%)[24]. This neglect could be in form of health workers not being present at the time of birth, not providing medications or not communicating the progress of labor. More than a quarter of women in this study responded that the health workers did not respond to their need whether or not asked, which is quite a large figure compared to those reported previously. The reason for not responding to needs could be the heavy workload of midwives and health care workers[25]. In an overburdened Nepali health care system where patient to healthcare workers ratio is unimaginably high [26]. In a communication with Shanti Kandel, RN (January 2021), six thousand delivery in Bheri Hospital is attended by a group of only eleven staffs for the fiscal year 2076/77. Silence can be a way in which a system defends itself against the many needs of patients. Silence from a care provider can cause neglect, resulting to avoidable complications during delivery [25], negative impact on the health of mother and / or baby [27] and also unwillingness to return or recommend others to the health facility for next delivery [28]. Respondents with complications are generally more likely to report D & A during delivery[29]. Although a third of respondents in this study had complications, rates of D & A remained low.

Women, during the process of delivery, are vulnerable to being abused by health workers whether it might be physical or verbal [28,30,31]. Such abuses are likely to result in a high rate of traumatic birth experience for women [32]. Different forms of abuse like being slapped (5.6%) or being shouted at (4.6%) has been reported in this study. Women experiencing physical and verbal abuse was found to be dramatically higher in another study conducted in central Nepal, which reported physical and verbal abuse to be 18.7% and 30% respectively [33]. The difference in reported abuse thus requires extensive research to identify the prevalence and institute appropriate interventions. A study has demonstrated that midwives feel a strong sense of accountability and responsibility for labor and delivery outcomes and tend to do whatever it takes to deliver a live baby to a healthy mother [34]. In addition, the midwives/ nurses ratio per population for Nepal is lesser (31.08/10,000 population) than the recommended by WHO (40/10,000 population) leading to overburden for health workers [35]. WHO recognizes that D & A not only violate the rights of women to respectful care, but also threaten their rights to life, health, bodily integrity and freedom from discrimination [36]. However, abuse in any form, whether it be physical or verbal should never be tolerated during labor and delivery.

Nearly half of the respondents (48.3 %) agreed that they were kept waiting for a long time, which is substantially higher than the study conducted in Ghana, Guinea, Myanmar, and Nigeria where 22% reported waiting for long periods before being attended by health workers [4]. The delay in care (27.8%) might be due to too few staff as compared to patients as revealed by a study of midwives of Malawi [8]. The situation might be similar in Nepal as shortage of staffs in the hospital setting has been reported [37], further worsened by the ongoing shifts in staff allocation as a part of Federalism.

Few participants i.e. approximately three out of a hundred, agreed that the health workers did not treat them well because of personal or their companions’ attributes, which is in contrast to a study conducted in Nigeria that showed a higher percentage of discrimination faced by respondents at 8.1% [38]. Birth preparedness practice in Nepal tends to be higher as reported by a study that denotes familiarization of pregnant women with the delivery setting. Early communication and interpersonal relationship between care provider and patient could be a cause for higher Discrimination-free Care [39].

Women with SVD were two times more likely to have a neutral response about RMC for Timely Care as compared to those who had cesarean delivery. Women experiencing caesarian delivery could have ultimately perceived the urgency of services for delivering a healthy baby, thus women with SVD would be more likely to report neutral Timely Care. Similarly, those who earn less than twenty thousand Nepalese Rupees were twice as likely to feel they had a high level of timely RMC. Women of lower economic status may be more tolerant of a long wait in order to receive care in a government facility with higher case load as opposed to delivering at home.

## Limitations

This study has used a standardized tool to gather quantitative information about D & A faced by women during labor and delivery. However in-depth insight regarding health workers’ perceptions and/or those of delivering women could not be obtained. Also, the potential for generalization of the findings cannot be ascertained as only a single tertiary care center in western Nepal has been included for the study.

Information bias and courtesy bias might have occurred as the information was collected by an on- duty student nurse, although the respondents were assured prior to the study that their opinion would have no impact on further treatment.

## Recommendations

Any forms of D & A must be prohibited during labor and delivery so that women can enjoy their experience of labor and delivery. Irrespective of the health system or staff-related issues; timely care must be of priority in order to ensure quality maternity care. Timely Care is simply not always possible in highly constrained settings such as Bheri Hospital, but the perception of Timely Care might possibly be reduced by additional communication and explanations of the cause of the delay.

Further studies should be conducted to determine RMC at all levels of the healthcare system throughout the country, such that potential generalization of our findings and appropriate interventions for improvement can be planned accordingly.

Sustained interaction with the health system are required to implement behavior change intervention central to promoting respectful care [40]. The successful improvement in maternity care environment for women and midwives needs broader interdisciplinary perspectives on the wider drivers of midwives’ disrespectful attitudes and behaviors [8].

## Conclusion

This study concludes that RMC is practiced highly in western Nepal in terms of Friendly Care, Abuse-free Care, and Discrimination-free Care. However, Timely Care is less reported.

Therefore, appropriate interventions to provide Timely Care to delivering women must be instituted. Along with this, adequate communication and explanation of delay can reduce perception of delayed care among care recipients. Physical or verbal abuse during labor and delivery must not be tolerated, and while rates of abuse were shown to be low in this study, there remains room for improvement. Further research on RMC in Nepal is required to clarify the drivers for D & A and examine potential solutions.

## Data Availability

Data will be made available upon request

## Abbreviations

D&A: Disrespect & Abuse
LSCS –: Lower Segment Caesarion Section
MMR: Maternal Mortality Ratio
NHRC: Nepal Health Research Council
RMC: Respectful Maternity Care
SDG –: Sustainable Development Goals
SVD –: Spontaneous Vaginal Delivery
WHO: World Health Organization

## Declaration

### Competing interests

The authors declare no competing interests.

### Funding

The study was self-funded by the authors.

## Acknowledgments

The authors are immensely thankful to the management of Bheri Hospital and to the NHRC for ethical approval. We express sincere gratitude to all women who participated in this study along with delivery room residents and midwives for providing support during the study. At last but not the least, we are thankful to all the student nurses of Sushma Koirala Memorial Hospital, Nepalgunj, Banke for their support during data collection.

## Notes

### Competing Interest Statement

The authors have declared no competing interest.

### Funding Statement

None

### Author Declarations

Ethical approval was obtained from the Nepal Health Research Council (Ref #1953, 18 March 2020). Written permission from Bheri Hospital administration was also obtained.

